# Early chest computed tomography to diagnose COVID-19 from suspected patients: A multicenter retrospective study

**DOI:** 10.1101/2020.03.24.20042432

**Authors:** Congliang Miao, Mengdi Jin, Li Miao, Xinying Yang, Peng Huang, Huanwen Xiong, Peijie Huang, Qi Zhao, Jiang Du, Jiang Hong

**Affiliations:** Department of Internal and Emergency Medicine, Shanghai General Hospital, Shanghai Jiao Tong University School of Medicine, Shanghai 201600, China; Department of Respiratory Diseases, High-tech Hospital, First Hospital Affiliated to Nanchang University, Jiangxi 330000, China; Department of Internal and Emergency Medicine, Shanghai General Hospital of Nanjing Medical University, Shanghai 201600, China; Department of Infectious Diseases, People’s Hospital of Yichun city, Jiangxi 336000, China

**Keywords:** Coronavirus infections, Viral pneumonia, Computed Tomography, Sensitivity and Specificity, Reverse-transcription polymerase-chain-reaction

## Abstract

**Objective:** The purpose of this study is to distinguish the imaging features of COVID-19 with other chest infectious diseases and evaluate diagnostic value of chest CT for suspected patients.

**Methods:** Adult suspected patients aged>18 years within 14 days who underwent chest CT scan and reverse-transcription polymerase-chain-reaction (RT-PCR) tests were enrolled. The enrolled patients were confirmed and grouped according to results of RT-PCR tests. The data of basic demographics, single chest CT features, and combined chest CT features were analyzed for confirmed and non-confirmed groups.

**Results:** A total of 130 patients were enrolled with 54 cases positive and 76 cases negative. The typical CT imaging features of positive group were ground glass opacity (GGO), crazy-paving pattern and air bronchogram. The lesions were mostly distributed bilaterally, close to the lower lungs or the pleura. When features combined, GGO with bilateral pulmonary distribution and GGO with pleural distribution were more common, of which were 31 cases (57.4%) and 30 cases (55.6%) respectively. The combinations were almost presented statistically significant (*P*<0.05) except for the combination of GGO with consolidation. Most combinations presented relatively low sensitivity but extremely high specificity. The average specificity of these combinations is around 90%.

**Conclusions:** The combinations of GGO could be useful in the identification and differential diagnosis of COVID-19, which alerts clinicians to isolate patients for treatment promptly and repeat RT-PCR tests until incubation ends.

## 1. Introduction

An outbreak of unexplained pneumonia has occurred in Wuhan City, Hubei Province, China, since December of 2019[1]. The virus causing the epidemic was detected as a new coronavirus (2019-nCoV), and the pneumonia caused was then named by the WHO as Corona Virus Disease 2019 (COVID-19)[2-4]. As to March 17, 2020, a total of 81118 patients in China were diagnosed as confirmed COVID-19, and 68802 patients have been cured and discharged[5]. At the same time, the number of suspected cases has also decreased significantly. It shows China’s epidemic prevention policy has achieved initial results. However, the infection has spread over 48 countries worldwide, especially in Iran, Korea and some European countries, even affected the United States. The global risk of this epidemic was escalated to highest level by WHO on February 28. Fast and effective diagnostics methods of this disease is of great importance at present.

According to the latest Diagnosis and Treatment Program for COVID-19 of China[6], the diagnosis of COVID-19 requires reverse-transcription polymerase-chain-reaction (RT-PCR) test, while a certain rate of false negative results has been reported [7]. Its limited production and relatively long testing period might not be conducive for screening. Accumulated clinical experience suggests that early chest computed tomography might be helpful in differential diagnosis of suspected cases. Recently, Chung, Fang and Kanne etc. had reported the chest CT imaging features of COVID19[8-10]. Fang and Ai further studied the sensitivity of Chest CT compared to RT-PCR with relatively small sample[9, 11]. However, there is no agreed standard for how to confirm a COVID-19 patient with these features. We retrospectively collected chest CT images of suspected pneumonia patients and divided them though RT-PCR test into positive and negative group, then compared the difference of CT features between two groups. Our purpose is to use statistics methods to distinguish the imaging features of COVID-19 with other chest infectious diseases and evaluate diagnostic value of chest CT for suspected patients.

## 2. Materials and Methods

### 2.1. Patient population

Data of this multicenter retrospective study were collected in three general hospitals of two provinces in China. We enrolled a continuous sample of suspected COVID-19 patients from January 12, 2020 to February 13, 2020 by the inclusion criteria as follows: (1) suspected patients of COVID-19 according to the 6^th^ edition of the Diagnosis and treatment program for COVID-19 of China [6]; (2) over 18 years old; (3) less than 14 days from onset to first CT. Exclusion criteria included: (1) past history with chronic lung disease; (2) pregnant women. Finally, 130 cases participated in our research. These patients’ general information, epidemiological history and CT imaging data were retrospectively collected.

### 2.2. Study design

The diagnostic criteria for suspected cases are as defined in the 6^th^ edition of the Diagnosis and treatment program for COVID-19 of China[6]. Nasopharyngeal swabs or sputum specimens were collected for RT-PCR tests. Patients with first negative results should receive repeated tests after an interval of at least 1 day. All suspected cases were divided into positive and negative groups according to the results of RT-PCR test. Patients who were negative before but positive on re-tests will eventually be defined as positive.

All patients were arranged to take CT scan examination, 33 patients were scanned on Optima 670 CT scanner, GE; 41 patients were scanned on Revolution Frontier, GE; 56 patients were scanned on SOMATOM Definition Flash, Japan. Examinations followed the normal chest protocols. Overall scan time was 2s, and slice thickness for reconstruction was 1.25 mm. All the reports were issued after double-blind reviews by two radiologists and would be concluded by a chief radiologist when opinions diverge.

This study was approved by the Ethics of Committees of local hospital. Informed consent for this retrospective study was waived.

### 2.3. Qualitative image analysis

According to the newly reported CT imaging features of COVID-19 and previous studies on [12], we summarized the performances that may appear on the early chest CT. Lesion morphology was described as presence of (1) ground-glass opacity (GGO); (2) consolidation; (3) crazy-paving pattern; (4) air bronchogram; (5) cavitation; (6) pulmonary nodule; (7) lymphadenopathy; (8) pleural effusion; (9) pulmonary atelectasis; (10) pleural thickening. Lesion distribution, such as whether they are unilateral, bilateral or peripleural, and the numbers of lung lobes and involvements of the upper, middle, and lower fields of lung were also counted.

### 2.4. Statistical analysis

The data of patients were recorded by Epidata and statistical analysis was performed on SPSS 13.0 (IBM Corporation). Normally distributed continuous variables were expressed as 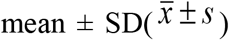 and skewed distribution was presented as the median (interquartile range). Independent t-test was used in comparison of the measurement data and Mann-Whitney U-test was used in it for skewed distribution. Calculators information are described by percentages and were compared using the chi-square test or Fisher method. All statistical tests were bidirectional comparison, and *P*<0.05 was regarded as statistically significant.

## 3. Results

### 3.1. Comparisons in basic demographics

A total of 166 patients were enrolled in this study. Excluding 2 cases younger than 18 years old, 28 cases with normal CT presentations, 2 cases with underlying lung disease, 2 cases with onset to consultation over 14 days, and 2 cases could not perform chest CT because of pregnancy, finally 130 cases were included in the statistical analysis with 54 cases positive and 76 cases negative (see Figure 1). As shown in Table 1, the mean age of positive group patients was 45.1 ± 13.4 years, while age ranged from 19 to 77 with 28 males (51.9%). The average time from symptoms onset to the first visit was 4 days. For the negative group patients, the average age was 41.8 ± 13.6 years, while age ranged from 19 to 81 with 49 males (64.5%). The average time from the onset of symptoms to the first visit was 3 days. It was found that proportion of Wuhan residence history and clustering incidence between two groups were statistically significant (*P*<0.05), while the remaining variables were not significant.

**Table 1.**
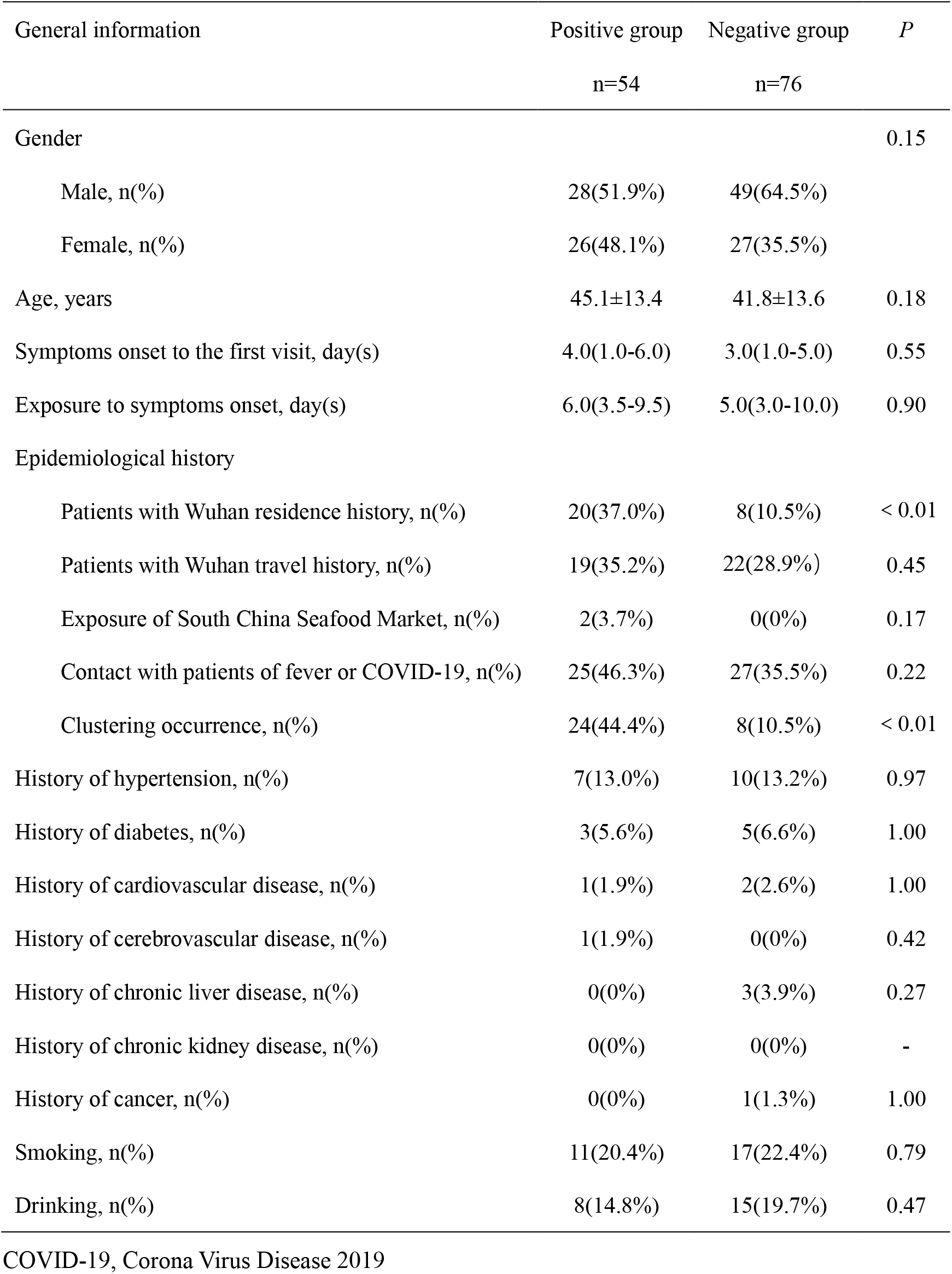
General demographic information.

| General information | Positive group<br>n=54 | Negative group<br>n=76 | *P* |
| --- | --- | --- | --- |
| Gender |  |  | 0.15 |
| Male, n(%) | 28(51.9%) | 49(64.5%) |  |
| Female, n(%) | 26(48.1%) | 27(35.5%) |  |
| Age, years | 45.1±13.4 | 41.8±13.6 | 0.18 |
| Symptoms onset to the first visit, day(s) | 4.0(1.0-6.0) | 3.0(1.0-5.0) | 0.55 |
| Exposure to symptoms onset, day(s) | 6.0(3.5-9.5) | 5.0(3.0-10.0) | 0.90 |
| Epidemiological history |  |  |  |
| Patients with Wuhan residence history, n(%) | 20(37.0%) | 8(10.5%) | < 0.01 |
| Patients with Wuhan travel history, n(%) | 19(35.2%) | 22(28.9%) | 0.45 |
| Exposure of South China Seafood Market, n(%) | 2(3.7%) | 0(0%) | 0.17 |
| Contact with patients of fever or COVID-19, n(%) | 25(46.3%) | 27(35.5%) | 0.22 |
| Clustering occurrence, n(%) | 24(44.4%) | 8(10.5%) | < 0.01 |
| History of hypertension, n(%) | 7(13.0%) | 10(13.2%) | 0.97 |
| History of diabetes, n(%) | 3(5.6%) | 5(6.6%) | 1.00 |
| History of cardiovascular disease, n(%) | 1(1.9%) | 2(2.6%) | 1.00 |
| History of cerebrovascular disease, n(%) | 1(1.9%) | 0(0%) | 0.42 |
| History of chronic liver disease, n(%) | 0(0%) | 3(3.9%) | 0.27 |
| History of chronic kidney disease, n(%) | 0(0%) | 0(0%) | - |
| History of cancer, n(%) | 0(0%) | 1(1.3%) | 1.00 |
| Smoking, n(%) | 11(20.4%) | 17(22.4%) | 0.79 |
| Drinking, n(%) | 8(14.8%) | 15(19.7%) | 0.47 |

COVID-19, Corona Virus Disease 2019

**Figure 1.**
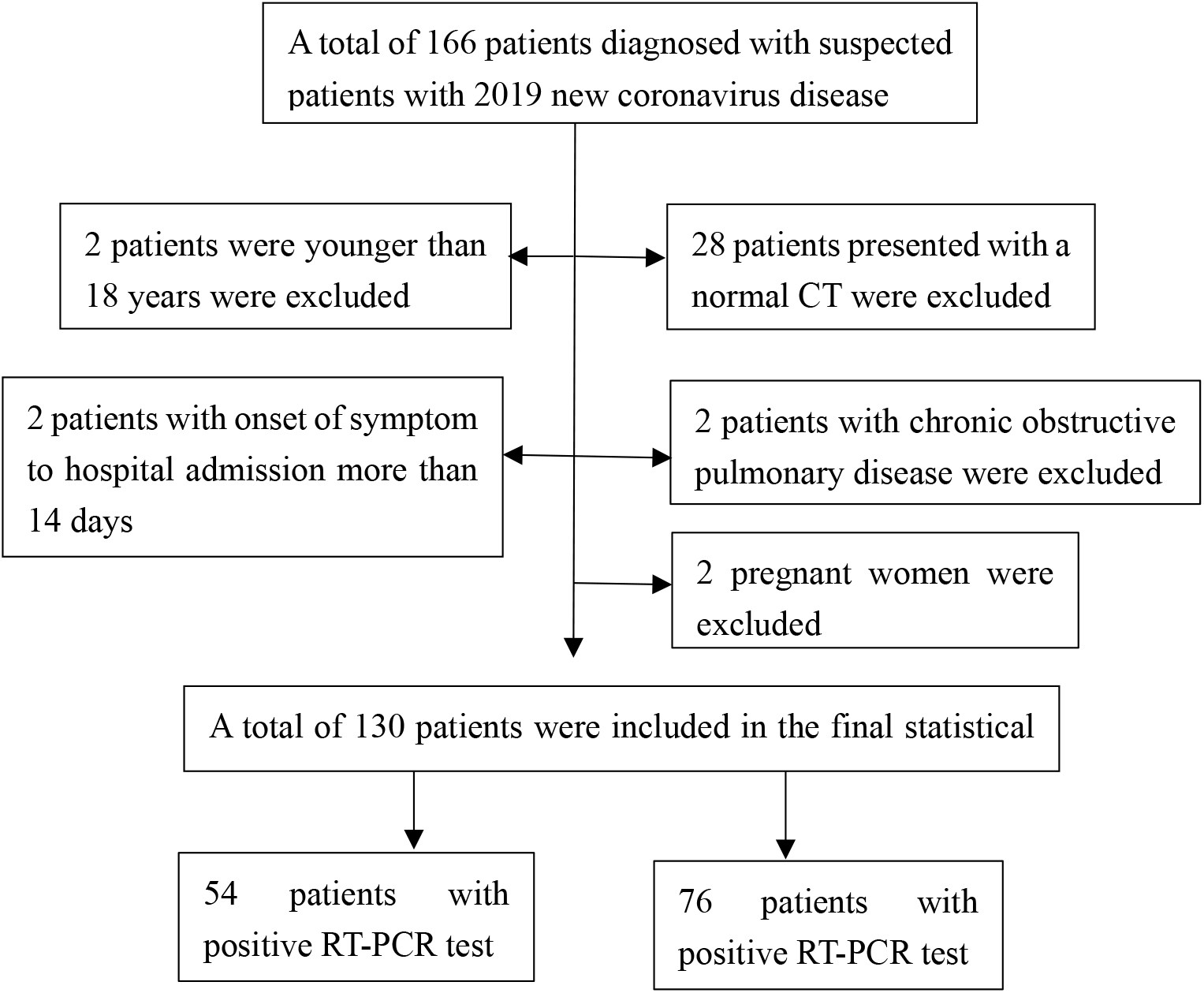
Flow chart of the study.

**Figure 2.**
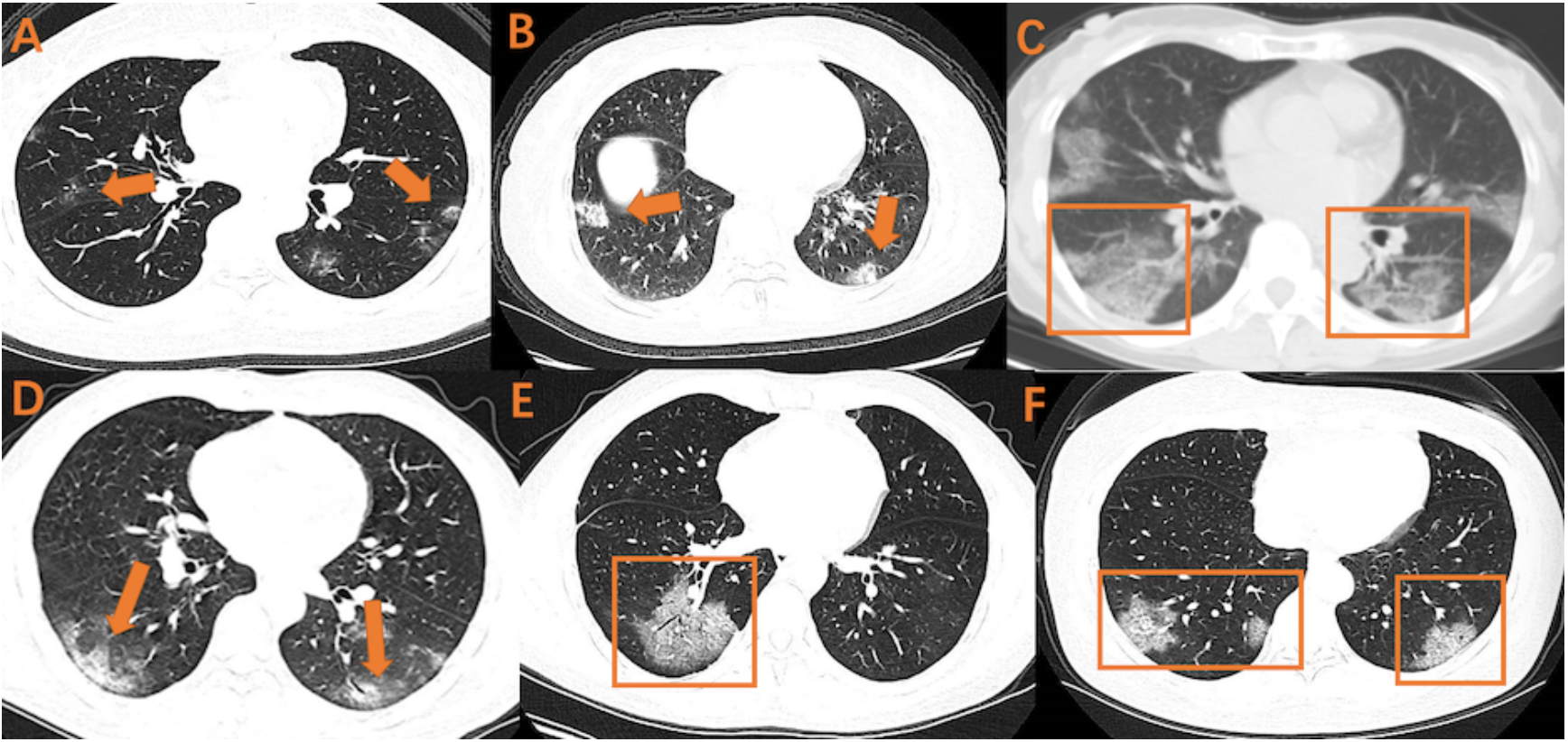
Various combinations of GGO on early CT imaging. GGO, ground glass opacity Various combinations of GGO on early CT imaging were shown as above. A, B, C: GGO with bilateral and subpleural distribution; D: GGO with air bronchogram and bilateral lower pulmonary distribution; E: GGO with air bronchogram; F: GGO with crazy-paving pattern and bilateral lower pulmonary distribution.

### 3.2. Comparisons in single CT imaging feature

Lesion morphology is shown in Table 2. Positive group patients in early stage mostly had GGO (38 cases, 70.4%), a typical imaging manifestation in viral pneumonia, followed by crazy-paving pattern (16 cases, 29.6%) and air bronchogram (14 cases, 25.9%), then consolidation (12 cases, 22.2%) and pleural thickening (11 cases, 20.4%). Pulmonary nodule, pleural effusion, lymphadenopathy and pulmonary atelectasis were rare. GGO, crazy-paving pattern, air bronchogram, and pleural thickening were proved to be statistically significant (*P*<0.05). The remaining variables were not (*P*>0.05).

**Table 2.** Lesion morphology of early chest CT.

| Morphological characteristics | Positive group | Negative group | *P* |
| --- | --- | --- | --- |
|  | n=54 | n=76 |  |
| GGO | 38(70.4%) | 32(42.1%) | 0.01 |
| Consolidation | 12(22.2%) | 29(38.2%) | 0.05 |
| Crazy-paving pattern | 16(29.6%) | 3(3.9%) | < 0.01 |
| Air bronchogram | 14(25.9%) | 5(6.6%) | < 0.01 |
| Cavitation | 1(1.9%) | 1(1.3%) | 1.00 |
| Pulmonary nodule | 8(14.8%) | 15(19.7%) | 0.47 |
| Lymphadenopathy | 4(7.4%) | 0(0%) | 0.03 |
| Pleural effusion | 0(0%) | 4(5.3%) | 0.14 |
| Pulmonary atelectasis | 1(1.9%) | 2(2.6%) | 1.00 |
| Pleural thickening | 11(20.4%) | 4(5.3%) | < 0.01 |

GGO, ground glass opacity.

Lesion distribution characteristics are showed in Table 3. For positive group, lesions in early stage were mainly distributed bilaterally (40 cases, 74.1%) and near the pleura (33 cases, 61.1%). Two lobes and all lobes involved were more common, with 18 cases (33.3%) and 13 cases (24.1%) respectively. 45 cases (83.3%) involving at least two lobes. Left lower lobe (40, 74.1%) and the right lower lobe (43, 79.6%) involvements were also frequently seen. Peripheral, bilateral or lower lung distribution and multi-lobe involvement proved to be statistically significant (*P*<0.05), and the remaining variables were not significant (*P*>0.05).

**Table 3.** Lesion distribution of early chest CT.

| Distribution characteristics | Positive group<br>n=54 | Negative group<br>n=76 | *P* |
| --- | --- | --- | --- |
| Bilateral pulmonary distribution | 40(74.1%) | 30(40.8%) | < 0.01 |
| Bilateral lower pulmonary distribution | 32(59.3%) | 19(25.0%) | < 0.01 |
| Peripleural distribution | 33(61.1%) | 21(27.6%) | < 0.01 |
| Distribution of lung lobes |  |  |  |
| Right upper lobe | 26(48.1%) | 27(35.5%) | 0.15 |
| Right middle lobe | 22(40.7%) | 23(30.3%) | 0.22 |
| Right lower lobe | 43(79.6%) | 35(46.1%) | < 0.01 |
| Left upper lobe | 28(51.9%) | 31(40.8%) | 0.21 |
| Left lower lobe | 40(74.1%) | 37(48.7%) | < 0.01 |
| Number of affected lobes |  |  | < 0.01 |
| Patients of 1 affected lobe | 9(16.7%) | 37(48.7%) |  |
| Patients of 2 affected lobes | 18(33.3%) | 15(19.7%) |  |
| Patients of 3 affected lobes | 7(13.0%) | 13(17.1%) |  |
| Patients of 4 affected lobes | 7(13.0%) | 1(1.3%) |  |
| Patients of 5 affected lobes | 13(24.1%) | 10(13.2%) |  |
| Patients of more than 2 affected lobes | 45(83.3%) | 39(51.3%) | < 0.01 |

### 3.3 Comparisons of combined early CT imaging features

According to the statistical results in Tables 2 and 3, we found that GGO was the basic manifestation in the positive group. To increase distinguishability on imaging, we combined GGO with other statistically significant features. Table 4 shows the various combinations of imaging features on early CT. When two features combined, it showed that the combination of GGO with bilateral pulmonary distribution, also the combination of GGO with pleural distribution were more common, of which were 31 cases (57.4%) and 30 cases (55.6%) respectively. When three items combined, 26 cases (48.1%) were presented as GGO with bilateral pulmonary and peripleural distribution. It presented the combination of GGO, bilateral pulmonary distribution, crazy-paving pattern, pleura distribution was the most with 12 cases (22.2%) when four items combined. They were almost presented statistically significant (*P*<0.05) except for the combination of GGO with consolidation.

**Table 4.** Combined early CT imaging features between positive group and negative group.

| Combined features | Positive group<br>n=54 | Negative group<br>n=76 | *P* |
| --- | --- | --- | --- |
| GGO+Consolidation | 11(20.4%) | 8(10.5%) | 0.12 |
| GGO+ Crazy-paving pattern | 16(29.6%) | 3(3.9%) | < 0.01 |
| GGO+Air bronchogram | 14(15.9%) | 4(5.3%) | < 0.01 |
| GGO+Bilateral pulmonary distribution | 31(57.4%) | 15(19.7%) | < 0.01 |
| GGO+Bilateral lower pulmonary<br>distribution | 21(38.9%) | 8(10.5%) | < 0.01 |
| GGO+Peripleural distribution | 30(55.6%) | 16(21.1%) | < 0.01 |
| GGO+Bilateral lower pulmonary<br>distribution+Peripleural distribution | 17(31.5%) | 5(6.6%) | < 0.01 |
| GGO+Bilateral pulmonary<br>distribution+Peripleural distribution | 26(48.1%) | 8(10.5%) | < 0.01 |
| GGO+Crazy-paving pattern+Peripleural<br>distribution | 12(22.2%) | 3(3.9%) | < 0.01 |
| GGO+Air bronchogram +Peripleural<br>distribution | 11(20.4%) | 2(2.6%) | < 0.01 |
| GGO+Crazy-paving pattern+Bilateral<br>lower pulmonary distribution | 11(20.4%) | 1(1.3%) | < 0.01 |
| GGO+Air bronchogram+Bilateral lower<br>pulmonary distribution | 8(14.8%) | 1(1.3%) | < 0.01 |
| GGO+Bilateral pulmonary<br>distribution+Crazy-paving Pattern+ Pleural<br>distribution | 12(22.2%) | 2(2.6%) | < 0.01 |
| GGO+bilateral pulmonary distribution+Air<br>bronchogram+ Peripleural distribution | 10(18.5%) | 1(1.3%) | < 0.01 |
| GGO+Bilateral lower pulmonary<br>distribution+Crazy-paving<br>pattern+Peripleural distribution | 9(16.7%) | 1(1.3%) | < 0.01 |
| GGO+Bilateral lower pulmonary<br>distribution+Air bronchogram +Peripleural<br>distribution | 6(11.1%) | 1(1.3%) | 0.02 |

GGO, ground glass opacity

### 3.4. Comparisons of diagnostic value of combined imaging features between the positive and negative groups

As shown on Table 5, the GGO with bilateral pulmonary distribution and GGO with peripleural distribution both presented highest as 57% on sensitivity, followed by GGO combined with bilateral pulmonary distribution and peripleural distribution as at 48%. The remaining combinations were around 25%. Contrarily, the significance of combination presented more obvious on specificity. When combining two items, the specificity of GGO with crazy-paving pattern was up to 96%, followed by GGO with air bronchogram (95%). When three features were combined, the specificity was over 95%, and GGO with crazy-paving pattern and bilateral pulmonary distribution was the highest at 99%. Specificity of diagnosis was almost 99% for four items.

**Table 5.** The diagnostic value of combined early CT imaging features between positive group and negative group.

| Combined features | Sensitivity | Specificity |
| --- | --- | --- |
| GGO+ Crazy-paving pattern | 0.30 | 0.96 |
| GGO+Air bronchogram | 0.26 | 0.95 |
| GGO+Bilateral pulmonary distribution | 0.57 | 0.81 |
| GGO+Bilateral lower pulmonary distribution | 0.39 | 0.89 |
| GGO+Peripleural distribution | 0.57 | 0.79 |
| GGO+Bilateral lower pulmonary distribution+Peripleural distribution | 0.31 | 0.93 |
| GGO+Bilateral pulmonary distribution+Peripleural distribution | 0.48 | 0.89 |
| GGO+Crazy-paving pattern+Peripleural distribution | 0.22 | 0.96 |
| GGO+Air bronchogram +Peripleural distribution | 0.20 | 0.97 |
| GGO+Crazy-paving pattern+Bilateral lower pulmonary distribution | 0.20 | 0.99 |
| GGO+Air bronchogram+Bilateral lower pulmonary distribution | 0.15 | 0.99 |
| GGO+Bilateral pulmonary distribution+Crazy-paving Pattern+ Peripleural distribution | 0.22 | 0.97 |
| GGO+bilateral pulmonary distribution+Air bronchogram+ Peripleural distribution | 0.19 | 0.99 |
| GGO+Bilateral lower pulmonary distribution+Crazy-paving pattern+Peripleural distribution | 0.17 | 0.99 |
| GGO+Bilateral lower pulmonary distribution+Air bronchogram +Peripleural distribution | 0.11 | 0.99 |

GGO, ground glass opacity

## 4. Discussion

In positive group, our study shows that GGO is the basic manifestation in CT imaging of COVID-19, followed by consolidation, crazy-paving pattern and air bronchogram. Lesions were mainly distributed bilaterally, close to lower lungs or the pleura. These findings are similar to those in previous descriptive researches on COVID-19[8, 10, 13, 14], which also overlapped with imaging of other viral pneumonias. For reason that we estimated pathological mechanism of COVID-19 as it might transmit though respiratory tract and damage terminal bronchioles and lung parenchyma near bronchiolar in the early stages, then in turn affect the entire lung lobules and diffuse alveolar damage like other viral pneumonias[15]. Apparently, it has more resemblances to the imaging of SARS (severe acute respiratory syndrome) and MERS (Middle East respiratory syndrome), which also belongs to coronavirus[16-19].

However, some distinguishable imaging features can be investigated as follows. The CT imaging features of SARS are mainly manifested by GGO with consolidation[20]. Lower lung distribution and peripheral involvement are more common, also more unifocal involvement than multifocal and bilateral involvement. While the manifestations of MERS on chest CT are mainly subpleural distribution, accompanied by extensive GGO and consolidation[16]. In addition, influenza virus pneumonia have high frequency of occurrence in winter and spring, with the manifestations of cavity, GGO and lobular distribution on chest CT[12].

Furtherly, we found that GGO, crazy-paving pattern, air bronchogram, and pleural thickening presented more on patients of COVID-19 than other viral pneumonias in chest CT imaging. On distribution, peripheral, bilateral or lower lung distribution and multi-lobe involvement also presented more on COVID-19. This may help us identify COVID-19 patients with chest CT scan. Previous researches have merely described the dynamic changes in chest CT imaging of COVID-19 and presented the features in the diagnosis so far[8, 10, 13, 14, 21]. Some of them had focused on the sensitivity compared to RT-PCR with relatively small sample size, while they did not compare these features with other viral pneumonias and explain how differential diagnosis was made by chest CT[9, 11]. Our study includes all suspected cases, including the positive and the negative, which may more useful for early differential diagnosis.

Theoretically, a definite diagnosis cannot be achieved on the basis of one single imaging feature, as it may overlap in imaging performances with other viral pneumonias as mentioned above. That’s why we then combined several statistically significant imaging features to evaluate the diagnostic value of chest CT scan. Apparently, when features combined, it presented relatively low sensitivity but extremely high specificity. The average specificity of all combinations is around 90%, and the combination of GGO with crazy-paving pattern and bilateral pulmonary distribution was the highest at 99% when three items combined. Even all reached to 99% for four items. Conclusion comes out that the combinations of GGO with different features in chest CT might significantly increase the specificity in diagnosis of COVID-19. It means in series of crazy-paving pattern, air bronchogram, bilateral lower pulmonary distribution, bilateral lower pulmonary distribution and pleural distribution, as long as they appear simultaneously with GGO on chest CT, COVID-19 should be highly suspected, even if the patient had repeatedly negative RT-PCR outcomes. This might help emergency physicians identify COVID-19 patients faster and more effectively. Based on accumulated experience, we recommend these patients to be quarantined and repeat the RT-PCR tests until the average incubation ends, especially in the areas with severe outbreaks. Worth noting that, COVID-19 is currently in the period of outbreak and as is still the season of all kinds of influenzas. Epidemiological exposure, RT-PCR test and chest CT scan are equally critical to diagnosis.

There are several limitations in our study. First, the method is to compare the difference of CT imaging features between confirmed patients and non-confirmed patients of COVID-19. Some early lesions might not be visible on chest CT since the maximum incubation period is 14 days or even more[6]. So the expression of time may not be precise enough according to the development of this disease. Second, the current gold standard for the diagnosis of COVID-19 is merely RT-PCR test[6]. Since the test samples are mostly pharynx swabs rather than bronchoalveolar lavage fluid (BALF), it might still present false negative results after repetitions.

In summary, the manifestations of COVID-19 vary. Some patients show similar manifestations with common viral pneumonias. However, it still has specific imaging characteristics. Moreover, the combinations of GGO could be useful in the identification and differential diagnosis of COVID-19, which alerts clinicians to isolate patients for treatment promptly and repeat RT-PCR tests until incubation ends.

## Data Availability

All data of this manuscript is available.

## Acknowledgements

None.

## Author contributions

The conception and design of the study: Jiang Du, Jiang Hong

Acquisition of data: Congliang Miao, Li Miao, Peng Huang, Huanwen Xiong,

Drafting the article: Mengdi Jin, Xinying Yang

Revising draft critically for important intellectual contents: Peijie Huang, Qi Zhao

Final approval of the version: Jiang Hong

## Abbreviations

2019-nCoV: 2019 novel coronavirus
COVID-19: Corona Virus Disease 2019
RT-PCR: reverse-transcription polymerase-chain-reaction
HRCT: High-resolution computed tomography
GGO: Ground glass opacity
MERS: Middle East respiratory syndrome
SARS: Severe acute respiratory syndrome
BALF: Bronchoalveolar lavage fluid

## References

[1] Lu R, Zhao X, Li J, Niu P, Yang B, Wu H, Wang W, Song H, Huang B, Zhu N et al: Genomic characterisation and epidemiology of 2019 novel coronavirus: implications for virus origins and receptor binding. Lancet 2020.

[2] Lu H, Stratton CW, Tang YW: Outbreak of pneumonia of unknown etiology in Wuhan, China: The mystery and the miracle. J Med Virol 2020, 92(4):401–402.

[3] Report of clustering pneumonia of unknown etiology in Wuhan City [http://wjw.wuhan.gov.cn/front/web/showDetail/2019123108989]

[4] Zhu N ZD, Wang W, et al.: A Novel Coronavirus from Patients with Pneumonia in China, 2019. NEJM 2020.

[5] Update on the outbreak of new coronavirus pneumonia as of 24:00 on March 17 [http://www.nhc.gov.cn/xcs/yqfkdt/202002/d5e15557ee534fcbb5aaa9301ea5235f.shtml]

[6] Diagnosis and treatment program for COVID-19(Trial version of the 7th Edition) [http://www.nhc.gov.cn/yzygj/s7653p/202002/8334a8326dd94d329df351d7da8aefc2/files/b21BQZKqdp2CV3QV5nUEsqSg1ygegLmqRygj.pdf]

[7] Xie X, Zhong Z, Zhao W, Zheng C, Wang F, Liu J: Chest CT for Typical 2019-nCoV Pneumonia: Relationship to Negative RT-PCR Testing. Radiology 2020:200343.

[8] Chung M, Bernheim A, Mei X, Zhang N, Huang M, Zeng X, Cui J, Xu W, Yang Y, Fayad ZA et al: CT Imaging Features of 2019 Novel Coronavirus (2019-nCoV). Radiology 2020:200230.

[9] Fang Y, Zhang H, Xie J, Lin M, Ying L, Pang P, Ji W: Sensitivity of Chest CT for COVID-19: Comparison to RT-PCR. Radiology 2020:200432.

[10] Kanne JP: Chest CT Findings in 2019 Novel Coronavirus (2019-nCoV) Infections from Wuhan, China: Key Points for the Radiologist. Radiology 2020:200241.

[11] Ai T, Yang Z, Hou H, Zhan C, Chen C, Lv W, Tao Q, Sun Z, Xia L: Correlation of Chest CT and RT-PCR Testing in Coronavirus Disease 2019 (COVID-19) in China: A Report of 1014 Cases. Radiology 2020:200642.

[12] Kim EA, Lee KS, Primack SL, Yoon HK, Byun HS, Kim TS, Suh GY, Kwon OJ, Han J: Viral pneumonias in adults: radiologic and pathologic findings. Radiographics 2002, 22 Spec No:S137–149.

[13] Lei J, Li J, Li X, Qi X: CT Imaging of the 2019 Novel Coronavirus (2019-nCoV) Pneumonia. Radiology 2020:200236.

[14] Shi H, Han X, Zheng C: Evolution of CT Manifestations in a Patient Recovered from 2019 Novel Coronavirus (2019-nCoV) Pneumonia in Wuhan, China. Radiology 2020:200269.

[15] Xu Z, Shi L, Wang Y, Zhang J, Huang L, Zhang C, Liu S, Zhao P, Liu H, Zhu L et al: Pathological findings of COVID-19 associated with acute respiratory distress syndrome. Lancet Respir Med 2020.

[16] Ajlan AM, Ahyad RA, Jamjoom LG, Alharthy A, Madani TA: Middle East respiratory syndrome coronavirus (MERS-CoV) infection: chest CT findings. AJR Am J Roentgenol 2014, 203(4):782–787.

[17] Das KM, Lee EY, Langer RD, Larsson SG: Middle East Respiratory Syndrome Coronavirus: What Does a Radiologist Need to Know? AJR Am J Roentgenol 2016, 206(6):1193–1201.

[18] Wong KT, Antonio GE, Hui DS, Lee N, Yuen EH, Wu A, Leung CB, Rainer TH, Cameron P, Chung SS et al: Thin-section CT of severe acute respiratory syndrome: evaluation of 73 patients exposed to or with the disease. Radiology 2003, 228(2):395–400.

[19] Zhao D, Ma D, Wang W, Wu H, Yuan C, Jia C, He W, Liu C, Chen J: Early X-ray and CT appearances of severe acute respiratory syndrome: an analysis of 28 cases. Chin Med J (Engl) 2003, 116(6):823–826.

[20] Koo HJ, Lim S, Choe J, Choi SH, Sung H, Do KH: Radiographic and CT Features of Viral Pneumonia. Radiographics 2018, 38(3):719–739.

[21] Pan F, Ye T, Sun P, Gui S, Liang B, Li L, Zheng D, Wang J, Hesketh RL, Yang L et al: Time Course of Lung Changes On Chest CT During Recovery From 2019 Novel Coronavirus (COVID-19) Pneumonia. Radiology 2020:200370.

